# Asymptomatic Employee Screening for SARS-CoV-2: Implementation of and Reactions to an Employer-Based Testing Program

**DOI:** 10.1101/2020.11.06.20227314

**Authors:** Laura H. Goetz, Tyler L. DeLaughder, Kathleen L. Kennedy, Nicholas J. Schork, Timothy K. McDaniel, Jeffrey M. Trent, David M. Engelthaler

**Author notes:** **Address correspondence to:** Laura H. Goetz, MD, MPH, The Translational Genomics Research Institute (TGen), 445 North Fifth Street, Phoenix, AZ 85004.

## Abstract

**Introduction:** Asymptomatic testing for SARS-CoV-2 among healthcare workers or other essential personnel could remove infected carriers from the workforce, decreasing chances for transmission and workplace outbreaks. Results from one-time testing programs have been reported but data regarding longitudinal testing, including information about employee’s reactions to such programs, is not readily available.

**Methods:** To identify asymptomatic carriers of SARS-CoV-2, we implemented a longitudinal screening program for critical on-site employees within our research institute in early April 2020. We conducted a survey of both on-site employees and those working from home in order to measure their reactions to the testing program. Statistical analysis of the survey was conducted with general linear regression and Pearson’s Chi-Square tests.

**Results:** Despite an ongoing high community prevalence rate of COVID-19, to date only two asymptomatic employees tested positive out of 1050 tests run during 7 months of the program. However, 12 symptomatic employees not participating in the program have tested positive. The employee survey was completed by 132/306 (43%) employees, with 93% agreeing that asymptomatic employee screening led to a better and safer working environment and 75% agreeing with on-site public health measures to help contain the virus, but only 58% feeling COVID-19 was a serious threat to their health.

**Conclusion:** Our results suggest that a longitudinal asymptomatic employee screening program for SARS-CoV-2 can be accepted by employees and can be used to maintain the health of the workforce, potentially keeping positivity rates below community levels in the face of the ongoing COVID-19 pandemic.

## INTRODUCTION

In January and February of 2020, the early months of the COVID-19 pandemic, reports from China and subsequently Italy regarding the high mortality rate of infected healthcare workers prompted consideration of stronger measures to keep healthcare workers from getting infected.^1,2^ Estimates of viral shedding and transmissibility indicated that peak viral loads could occur in many individuals prior to symptom onset, which was different from what was experienced in prior SARS coronavirus epidemics.^3-5^ Up to 68% of cases in some areas were thought to be due to pre-symptomatic transmission of SARS-CoV-2.^6-10^ Thus, with asymptomatic carriers identified as a significant source of disease spread, routine testing of asymptomatic individuals, followed by isolation of those testing positive and quarantining of their close contacts, emerged in many countries as an effective first step in the containment of outbreaks and flattening of the epidemiologic curve for COVID-19.^11-14^

In early March of 2020, as the pandemic spread throughout the United States, the Translational Genomics Research Institute’s (TGen’s) pathogen and microbiome facility was repurposed into a CLIA-registered COVID-19 clinical diagnostic laboratory, employing a SARS-CoV-2 RT-PCR assay, which was allowed under an Emergency Use Authorization program from the Federal Drug Administration (FDA). More than 45,000 suspect COVID-19 clinical samples were processed in the facility, with >5000 known positive specimens identified. Additionally, all positive specimens were genomically sequenced. The staff size in the facility increased from approximately 35 employees to over 70 at the height of the response, increasing chances for person-to-person spread from an unidentified asymptomatic infected employee.

Due to the potential high risk for workplace transmission through an increased employee population and presence of live virus in diagnostic samples, a longitudinal asymptomatic employee screening program was designed to conduct routine and repeated testing for SARS-CoV-2 infection, with subsequent isolation of individuals testing positive, in addition to contact tracing and at-home quarantine of identified close contacts, following CDC recommendations. The goal of this program was to decrease the risk for transmission of the virus in the workplace, primarily in order to maintain continual CLIA laboratory operations during the COVID-19 response. Table 1 lists potential benefits and barriers to such an employee screening program. Here we review our experience with development and implementation of this program, and present results to date, as well as results of an employee survey we conducted seven months after the program began.

**Table 1.** Possible benefits and barriers to implementation of an asymptomatic employee screening program for SARS-CoV-2.

| Benefits | Barriers |
| --- | --- |
| 1. Protecting health of the workforce by early identification of small outbreaks to reduce spread of SARS-CoV-2.<br>2. Minimizing workforce depletion due to unnecessary quarantine periods.<br>3. Building employer-employee trust and appreciation on both sides.<br>4. Psychological benefits of knowing public health practices are working to decrease spread of COVID-19. | 1. Element of coercion or fear of retribution for not participating.<br>2. Lack of interest in participation in the program.<br>3. Lack of trust that information will be kept confidential.<br>4. Fear of stigmatization if a positive result is leaked to co-workers.<br>5. Discomfort with the specimen collection process.<br>6. Fear of missing work if employee learned they were positive. |

## METHODS

### Program Description

After the lockdown and work-from-home orders were put into place in Arizona in late March of 2020, a protocol for longitudinal asymptomatic employee screening was designed to test the subset of employees working on-site as critical or essential workers in the SARS-CoV-2 testing facility or in research laboratories working with SARS-CoV-2. Those employees working full-time from home were not eligible to participate in the program. Symptomatic employees were offered testing outside of the asymptomatic employee screening program. Out of 306 total employees, 40 were initially eligible in April, increasing to 71 during July and August. The TGen Offices of Compliance, Human Resources and Legal Counsel approved the protocol and assisted in careful development of the consent forms to avoid any element of coercion, stressing that participation in the protocol was confidential and entirely voluntarily, and emphasizing that there would be no negative repercussions for not participating, such as loss of employment or reassignment of job duties.

Public health precautions followed in the workplace during March and April included daily symptom checking and temperature measurement upon arrival at work, social distancing, frequent hand-washing, and frequent disinfection of shared surfaces. While masks were always part of the required PPE in the laboratory, they became mandatory in all work areas starting in May when mask-wearing became part of the Center for Disease Control and Prevention workplace recommendations.^15^

An initial testing interval of two weeks was determined based on the community prevalence of COVID-19, which in early April was less than 20 per 100,000, and also on estimates of viral shedding and transmissibility in asymptomatic individuals.^3,4^ This interval was shortened to one week during spikes in community to prevalence to over 1,000 per 100,000 individuals.

### Employee Survey

Six months after implementation of the testing program, we developed a novel short IRB-exempt and completely anonymized Likert Type (“strongly agree” to “strongly disagree”) survey instrument to assess employee reactions to the program and the pandemic in general. No identifying information, such as sex and age, was collected. Five categories were assessed: feelings about the pandemic; beliefs about asymptomatic testing in general; feelings about an employee testing program; feelings about public health measures in the workplace, and reactions to the testing process itself.

The survey was developed as an online tool and an email invitation was sent to all employees describing the instrument and the reasoning behind it. Consent for participation was obtained when the individual agreed to begin the survey. Reminder emails were sent 1 and 2 weeks after the initial email. Statistical analysis of results included frequency measurements for work-from-home status, participation and motivation questions. Pearson’s Chi-square and generalized linear regression were used to assess associations between the items on the questionnaire and to ultimately test the hypothesis that employees working on site or participating in the program would differ in their responses compared with those working from home or not-participating. All analyses were carried out using R software, version 4.0.3.

## RESULTS

### Screening Program Results

The first screening session was held on April, 3, 2020 and will continue throughout the length of the pandemic. Thirty sessions were completed as of November 1, 2020. The average number of participants per session was 48, with a range of 11 to 71. The average participation rate of those eligible was 95% with a range of 85 to 100%. Of the 30 total sessions, 28 did not identify a single positive asymptomatic employee. To date, 1082 samples have been processed with only two asymptomatic positive employees identified. This equals a case rate of 184 per 100,000 individuals tested, which is far below the current case rate for Arizona of 3,253 per 100,000.^16^ One employee was retested due to concerns for a false positive and tested negative 4 days later. The other employee remained home for seven days, did not develop symptoms, was retested on day 7 and subsequently found to be negative.

### Symptomatic Testing Results

During the period from April to November 1, 2020, a total of 12 symptomatic employees not participating in asymptomatic screening tested positive for SARS-CoV-2. Nine of these employees were working on-site and three were working from home. Isolation, contact tracing, and quarantine of close contacts was conducted and no transmission to co-workers was identified. Employees working on-site were retested after they had been symptom-free for 72 hours and a minimum of seven days had passed since their last positive test. One individual remained positive for two weeks and the other 8 tested negative after 10 days. Since not all symptomatic employees who receive testing notify our Human Resources Department of their symptoms, the denominator for the total number of symptomatic employees tested is unknown.

### Survey Results

An initial email invitation was sent to a total of 306 employees, approximately half of whom worked from home full time and half of whom worked on-site either full time or part time during the COVID-19 pandemic response. Of those, 133 started and 132 completed the survey, for a response rate of 132/306 or 43%. Eighty three of the 132 (63%) respondents were working on-site and of these, 47 (57%) were participating in the asymptomatic screening program. Three of the 49 (6%) respondents working from home were participating in the asymptomatic employee testing program. Table 2 lists motivating factors for participation. Forty four of the 47 (94%) participating respondents chose item 2, “I wanted to know my status for my own benefit”, while 35 (75%) chose more than one of items 2 through 5 and 28 (60%) chose all four items 2 through 5. Only one individual chose item 1. Table 3 gives results for the each of the five categories of survey items. Regarding feelings about the pandemic, a small majority (57%) agreed or strongly agreed with the statement ‘COVID-19 is a serious threat to my personal health’, but over 70% felt more anxious since the pandemic. For items assessing beliefs about asymptomatic testing in general, 96% of employees felt that asymptomatic screening could help contain the epidemic, 93% felt that asymptomatic employee screening is necessary to keep the workforce healthy, and 89% felt this type of testing should be freely available to the public through the health department. Additionally, 86% did not feel asymptomatic screening should only be available for high-risk individuals. Regarding feelings specifically about the employee program, four individuals (3%) did not completely understand that the program was voluntary and 5 (4%) reported feeling pressured into participating in the program. Other findings of note included responses to questions concerning ‘feelings about public health measures in the workplace’, where the vast majority replied favorably regarding their perceptions of how their employer and coworkers followed basic guidelines to keep the workplace safe.

**Table 2.**
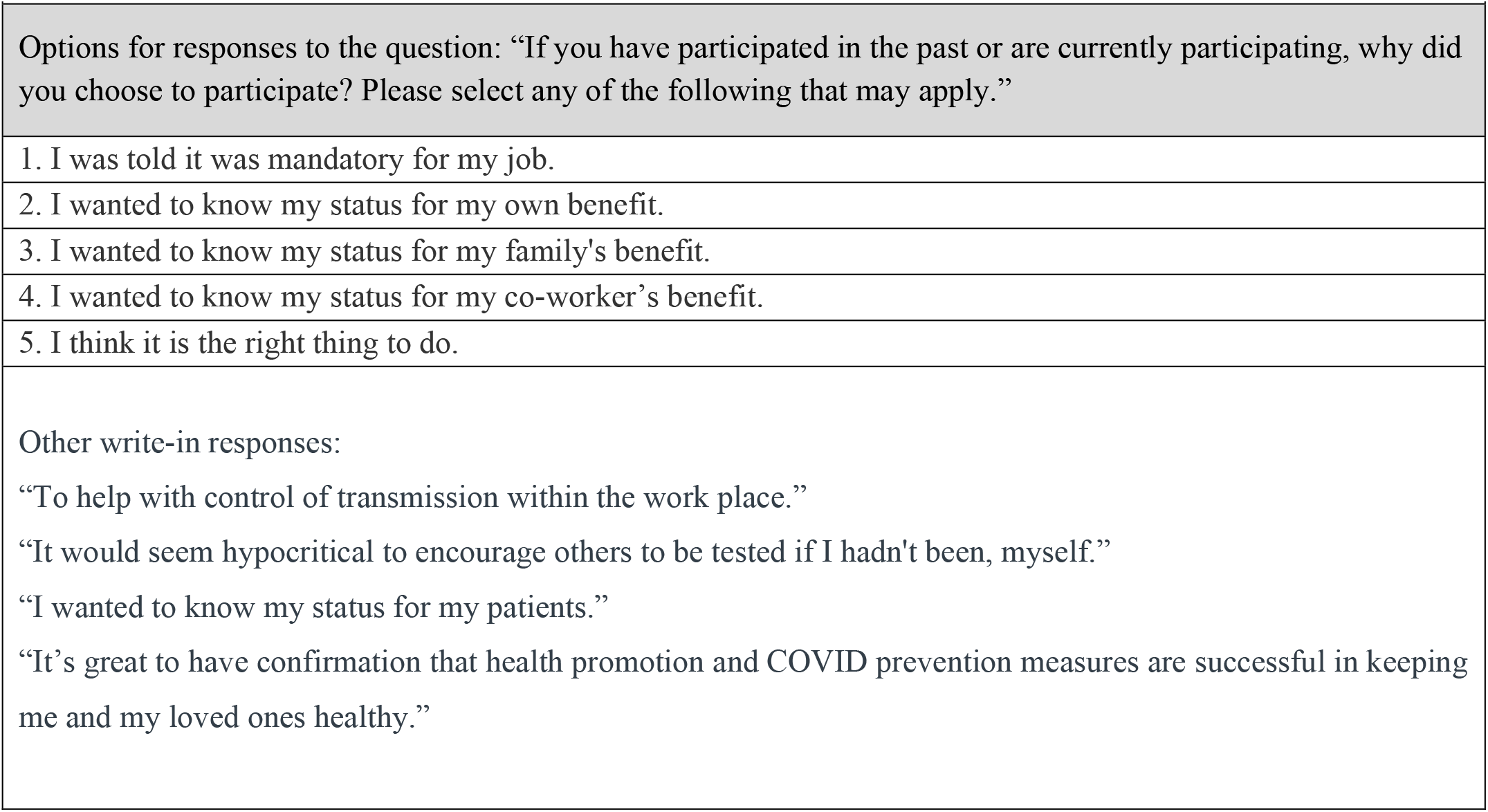
Motivations for participating in asymptomatic employee screening for SARS-CoV-2

**Table 3.**
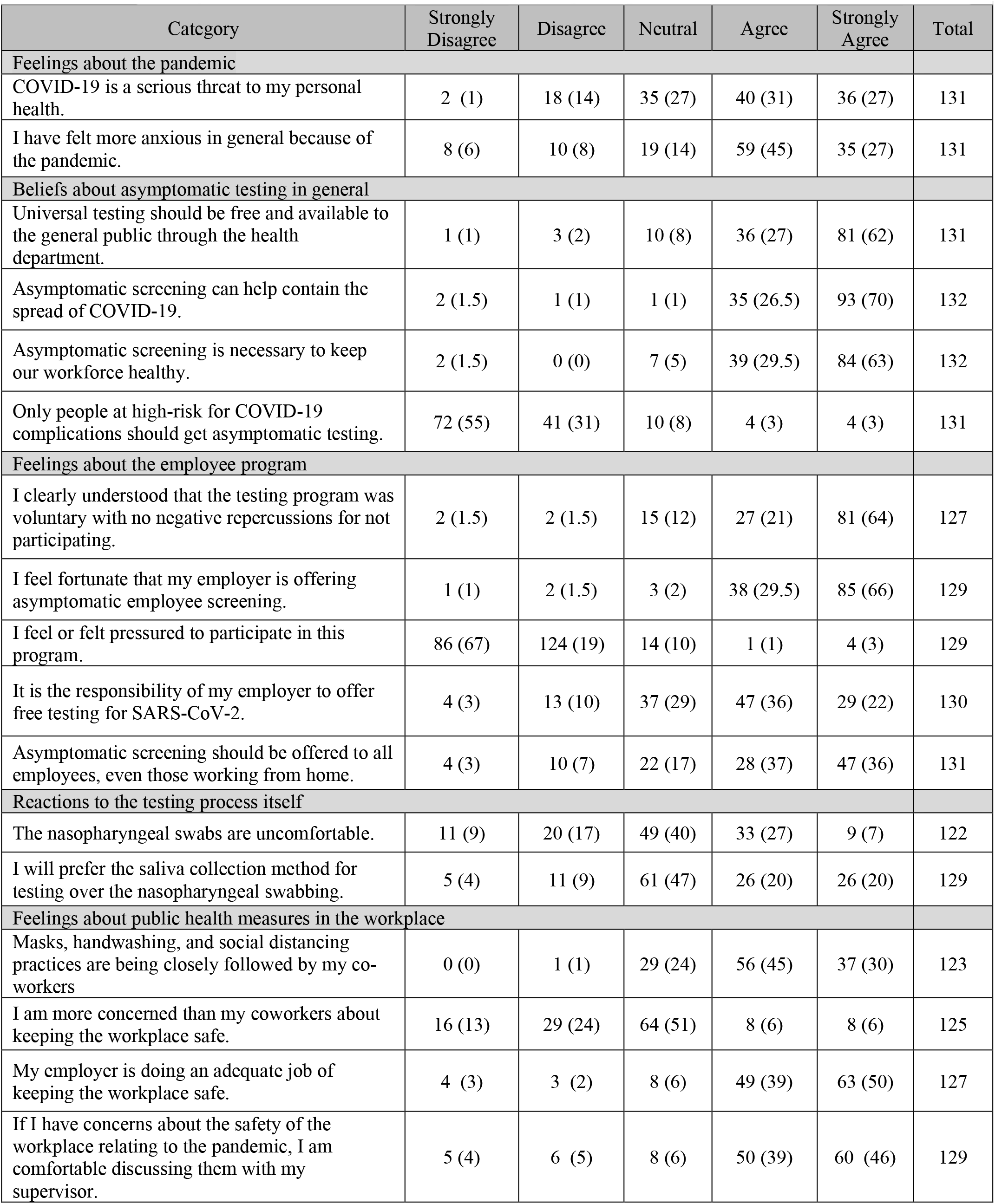
Selected survey results. Cells contain the number (percent) responding in each column.

Four individuals had participated in the past but stopped participating. Two of these individuals indicated the reason for stopping as “I did not feel my risk for getting COVID-19 was high enough for me to continue to get tested”; one individual chose “Masks, handwashing, and social distancing practices are enough to keep me safe from getting sick” and one individual chose “The testing process was inconvenient due to timing with work schedule, location, etc.”

Generalized linear regression analysis focusing on those working from home and those not working from home as a dependent variable did not suggest any significant differences in responses, nor between those participating and those not participating. We also used generalized linear regression with the question ‘COVID-19 is a serious threat to my personal health’ as the dependent variable and found no significant difference between responses from those working on site or not and those participating or not. Using Pearson’s Chi-square test on contingency tables reflecting employee responses, we assessed each survey item listed in Table 3 individually and found no significant difference in responses between non-participants and participants or those working from home or on-site.

## DISCUSSION

Since the COVID-19 pandemic first became widespread (Spring 2020), reports from around the world showed asymptomatic screening for SARS-CoV-2 to be an effective strategy for containing COVID-19 outbreaks and as such, we began a screening program for our employees in early April, 2020. Since that time, several groups have published recommendations for asymptomatic screening, either in a limited fashion for the healthcare workforce, such as Grassly et al^17^ and Black et al,^18^ or more broadly via universal testing, as described by McClellan et al^19^. The rationale put forth by these groups for such testing includes the early identification of small outbreaks to reduce spread, protecting the health of the workforce, and minimizing workforce depletion by unnecessary quarantine.

Several published reports have described either repeated or one-time screenings of asymptomatic healthcare workers and are summarized in Table 4. Jameson et al^20^ published their results of a voluntary one-time screening of healthcare workers in an urban hospital in Michigan during a time of high community burden of SARS-CoV-2, at 515 actives cases per 100,000. They reported a participation rate of 24.2% (121/499) of eligible employees, all of whom tested negative. This study provided the hospital with critical data showing that the precautionary measures they were using were effective. Nagler et al^21^ reported their results of on-demand testing of both asymptomatic and symptomatic employees of New York University’s Langone Health Center from March through May, 2020. They found the asymptomatic positivity proportion was 12% in March and decreased to 0% in May. They concluded that their program was effective in maintaining and managing the health of their workforce. Rivett et al^22^ reported their results from a one-time screening of asymptomatic healthcare workers over a 3 week period in April 2020 at Cambridge University’s National Health Service Hospital. They identified 30 positive individuals out of 1032 who were tested. These individuals were able to be removed from the workforce when they would have otherwise continued working. They concluded that their program showed the utility of routine screening of asymptomatic employees. Lombardi et al^23^ in Milan, Italy reported their results from one-time screening of at-risk healthcare workers at the Foundation IRCCS Ca’ Granda Ospedale Maggiore Policlinico who had contact with a known positive patient or co-worker during the peak of the pandemic in late February and March, 2020. They identified 17 positive out of the 1070 asymptomatic individuals who were tested (1.6%). Finally, Mohanty et al^24^ reported the results from a program that is most similar in design to the one implemented at our institute. They began asymptomatic screening of patients and staff in the electrophysiology (EP) units and Emergency Medical Services (EMS) of several hospitals in the United States. From April to June 2020, they tested 691 staff members, and identified 31 positive individuals (4.5%).They did not identify any new transmissions between coworkers or between staff and patients. They concluded the asymptomatic screening program led to a safe work environment with no evidence of hospital-acquired infection.

**Table 4.** Asymptomatic screening programs reported in the literature

| **Table 4.** Asymptomatic screening programs reported in the literature |  |  |  |  |
| --- | --- | --- | --- | --- |
| **Source** | **Employee Group** | **Screening Frequency** | **Number Screened** | **Asymptomatic Carrier Rate** |
| Jameson<sup>20</sup> | Michigan urban hospital workers | One time | 121 | 0.0% |
| Nagler<sup>21</sup> | New York City hospital workers | On-Demand over three months | 1596 | 12% in March and 0% in May |
| Rivett<sup>22</sup> | Cambridge, England hospital workers | One-time | 1302 | 2.3% |
| Lombardi<sup>23</sup> | Milan, Italy hospital workers | One-time | 1070 | 1.6% |
| Mohanty<sup>24</sup> | Electrophysiology Unit workers from multiple locations across the United States. | Repeat testing over 3 months | 691 | 4.5% |

Our testing results of an asymptomatic employee screening program in a COVID-19 diagnostic and research facility revealed an extremely low proportion of asymptomatic carriers, with just 2 individuals testing positive out of 1082 tests performed across seven months. Despite significant increases in the community case rate - the current case rate for Arizona is 3253 per 100,000^16^ - and the risks associated with being employed in a COVID-19 diagnostics and research laboratory, the infection rate amongst employees participating in asymptomatic screening has consistently been below expectation based on general community rates of infection and below rates of infection amongst employees not participating in the program. The low rate of infection in the screened group could be a reflection of a higher degree of adherence to public health measures such as handwashing and mask wearing. Whether or not participation in the screening program increased one’s likelihood of following public health measures is an interesting question, but not one we were able to capture with our survey instrument.

Notably, the employee survey identified strong beliefs in universal testing, following public health measures in the workplace and overwhelming support of asymptomatic employee screening, regardless of whether employees were working on-site or from home, or participating or not. One of the largest concerns when instituting the screening program was the potential perception among employees of coercion to participate. We attempted to address this by carefully drafting the consent forms for the testing program to clearly state that this program was entirely voluntary and confidential, and that there would be no repercussions for not participating. As seen from the survey results, less than 5% of people felt pressured into participating or felt that they did not clearly understand that the program was entirely voluntary, indicating our consenting process and communications about the program were effective at minimizing concerns of coercion.

Interestingly, only 58% of respondents felt that COVID-19 was a serious threat to their personal health. We did not find an association between perceptions of COVID-19 as a serious threat based on whether or not someone is working from home and/or participating in the program. Since participation in the program is entirely voluntary, our hypothesis was that individuals who are working from home and/or not participating do not necessarily feel COVID-19 is a threat, however this was not supported by our analyses. This could be due to the relatively small sample size.

There are several limitations to discuss. The number of employees participating in the screening program remains small, averaging less than 60 per week, and such a program may not be generalizable to larger workplaces. Additionally, the survey instrument we used has not yet been validated, although we plan on exploring this with future IRB-approved research. Our survey response rate was 43%, which is above reported averages for organizational surveys,^25^ but less than desired. Thus, the overwhelming support shown by respondents to the program could be due to volunteer bias, where those individuals who approve of such a program were more likely to complete the survey. Also, because of the need for complete anonymity we could not collect any demographic or identifying information, thus other important analyses of the survey results were not possible, such as whether or not the responses differed by sex or age of the participant. These variables are known to be important factors in understanding personal behaviors and beliefs about the pandemic ^26,27^ and by omitting them in our survey, generalizations to the broader population is limited. Another limitation is that we did not conduct a baseline survey prior to implementation of the program, so we are unable to assess whether the beliefs and attitudes about the testing program we collected post hoc were in fact altered by being part of the program, and not reflective of *a priori* beliefs and attitudes. In addition, the tested employee population was a group familiar with laboratory testing and medical research, so the results might not be applicable to a lay workforce.

## CONCLUSION

Despite the limitations listed above, we feel our results suggest that a longitudinal asymptomatic employee screening program for SARS-CoV-2 can be well received by employees and can help maintain the health of the workforce during the ongoing COVID-19 pandemic. We have learned that it is important to communicate clearly to address issues surrounding perceptions of employee coercion and to ensure that the voluntary nature of such a program is conveyed. We have also learned that extending the program to all employees, even those working from home, can possibly help decrease the spread of the pandemic in general. Finally, there is a role for expanding this type of screening program for other active respiratory infectious diseases, such as the influenza and respiratory syncytial viruses, since these can also be responsible for significant employee illness and absenteeism. The success of this program provides evidence for establishing similar programs even earlier in the face of future emergent public health threats.

## Data Availability

The data is available from the authors upon request.

## ACKNOWLEDGEMENTS

Aspects of this work were funded in part by philanthropic gifts from the Northern Arizona Regional Behavioral Health Authority (NARBHA) Institute and the Garcia Family Foundation. Dr. Laura Goetz contributed to the design, implementation and management of the testing program, created the employee survey, conducted the statistical analyses, and wrote the manuscript.

Dr. Tyler DeLaughder contributed to the design, implementation and management of the testing program and the employee survey and wrote parts of the manuscript.

Kathleen Kennedy is responsible for the employee health program, helped with conceptualization and design of the employee survey and helped revise and edit the manuscript.

Dr. Nicholas Schork advised on design and implementation of the employee testing program and all statistical analyses, and also revised and edited the manuscript.

Dr. Timothy McDaniel and Jeff Trent advised on the employee testing program and revised and edited the manuscript.

Dr. David Engelthaler oversaw the design and implementation of the employee testing program and revised and edited the manuscript.

We acknowledge Carmel Plude for carrying out the testing itself and contributing to the efficient running of the program.

## Financial disclosures

The authors have no financial disclosures or conflicts of interest.

## Notes

### Competing Interest Statement

The authors have declared no competing interest.

### Author Declarations

Western IRB was consulted and confirmed this study was exempt from IRB review.

## REFERENCES

1. Sadeghi N, Wen L. Novel Coronavirus Should Prompt Examination of Impact of Outbreaks on Health Care Workers. Health Affairs Blog https://wwwhealthaffairsorg/do/101377/hblog20200214587257/full/ Accessed March 24, 2020.

2. Stickings T, Dyer C. Five more Italian doctors die battling coronavirus: Thirteen medics have now lost their lives, with 2,629 health workers infected - 8.3% of country’s total. https://www.dailymail.co.uk/news/article-8129499/More-2-600-medical-workers-infected-coronavirus-Italy.html Accessed March 20, 2020.

3. Ganyani T, Kremer C, Chen D, et al. Estimating the generation interval for coronavirus disease (COVID-19) based on symptom onset data, March 2020. Euro Surveill. 2020;25(17).

4. He X, Lau EHY, Wu P, et al. Temporal dynamics in viral shedding and transmissibility of COVID-19. Nat Med. 2020;26(5):672–675.

5. Peiris JS, Chu CM, Cheng VC, et al. Clinical progression and viral load in a community outbreak of coronavirus-associated SARS pneumonia: a prospective study. Lancet. 2003;361(9371):1767–1772.

6. Mizumoto K, Kagaya K, Zarebski A, Chowell G. Estimating the asymptomatic proportion of coronavirus disease 2019 (COVID-19) cases on board the Diamond Princess cruise ship, Yokohama, Japan, 2020. Euro Surveill. 2020;25(10).

7. Qiu J. Covert coronavirus infections could be seeding new outbreaks. Nature. 2020.

8. Wu Z, McGoogan JM. Characteristics of and Important Lessons From the Coronavirus Disease 2019 (COVID-19) Outbreak in China: Summary of a Report of 72314 Cases From the Chinese Center for Disease Control and Prevention. JAMA. 2020;323(13):1239–1242.

9. Rothe C, Schunk M, Sothmann P, et al. Transmission of 2019-nCoV Infection from an Asymptomatic Contact in Germany. N Engl J Med. 2020;382(10):970–971.

10. Day M. Covid-19: four fifths of cases are asymptomatic, China figures indicate. BMJ. 2020;369:m1375.

11. Day M. Covid-19: identifying and isolating asymptomatic people helped eliminate virus in Italian village. BMJ. 2020;368:m1165.

12. McGregor G. South Korea amassed the world’s most comprehensive coronavirus data. What it’s taught us so far. https://fortune.com/2020/03/19/coronavirus-south-korea-test-data/ Accessed March 19, 2020.

13. Kaplan EH. Containing 2019-nCoV (Wuhan) coronavirus. Health Care Manag Sci. 2020;23(3):311–314.

14. Cristani A, Cassone A. In one Italian town, we showed mass testing could eradicate the coronavirus. https://www.theguardian.com/commentisfree/2020/mar/20/eradicated-coronavirus-mass-testing-covid-19-italy-vo Accessed March 24, 2020.

15. Center for Disease Control and Prevention. Interim U.S. Guidance for Risk Assessment and Work Restrictions for Healthcare Personnel with Potential Exposure to COVID-19 https://www.cdc.gov/coronavirus/2019-ncov/hcp/guidance-risk-assesment-hcp.html Accessed September 12, 2020.

16. Arizona Department of Health Services COVID-19 Data Dashboard https://azdhs.gov/preparedness/epidemiology-disease-control/infectious-disease-epidemiology/covid-19/dashboards/index.php Accessed November 1, 2020.

17. Grassly N, Pons-Salort M, Parker E, White P, al e. Report 16: Role of testing in COVID-19 control. https://ww.imperial.ac.uk/media/imperial-college/medicine/mrc-gida/2020-04-23-COVID19-Report-16.pdf Accessed September 12, 2020.

18. Black JRM, Bailey C, Przewrocka J, Dijkstra KK, Swanton C. COVID-19: the case for health-care worker screening to prevent hospital transmission. Lancet. 2020;395(10234):1418–1420.

19. McClellan M, Gottlieb S, Mostashari F, Rivers C, Silvis L. A National COVID-19 Surveillance System: Achieving Containment https://healthpolicy.duke.edu/sites/default/files/2020-06/a_national_covid_surveillance_system.pdf Accessed August 24th, 2020.

20. Jameson AP, Biersack MP, Sebastian TM, Jacques LR. SARS-CoV-2 screening of asymptomatic healthcare workers. Infect Control Hosp Epidemiol. 2020:1–2.

21. Nagler AR, Goldberg ER, Aguero-Rosenfeld ME, et al. Early Results from SARS-CoV-2 PCR testing of Healthcare Workers at an Academic Medical Center in New York City. Clin Infect Dis. 2020.

22. Rivett L, Sridhar S, Sparkes D, et al. Screening of healthcare workers for SARS-CoV-2 highlights the role of asymptomatic carriage in COVID-19 transmission. Elife. 2020;9.

23. Lombardi A, Consonni D, Carugno M, et al. Characteristics of 1573 healthcare workers who underwent nasopharyngeal swab testing for SARS-CoV-2 in Milan, Lombardy, Italy. Clin Microbiol Infect. 2020;26(10):1413 e1419–1413 e1413.

24. Mohanty S, Lakkireddy D, Trivedi C, et al. Creating a safe workplace by universal testing of SARS-CoV-2 infection in asymptomatic patients and healthcare workers in the electrophysiology units: a multi-center experience. J Interv Card Electrophysiol. 2020.

25. Baruch Y, Holtom B. Survey response rate levels and trends in organizational research. Human Relations. 2008;61(8):1139–1160.

26. Clements JM. Knowledge and Behaviors Toward COVID-19 Among US Residents During the Early Days of the Pandemic: Cross-Sectional Online Questionnaire. JMIR Public Health Surveill. 2020;6(2):e19161.

27. Fan Y, Orhun AYi, Turjeman D. Heterogeneous Actions, Beliefs, Constraints And Risk Tolerance During The Covid-19 Pandemic National Bureau Of Economic Research Working Paper Series Working Paper No 27211 May 2020 DOI: 103386/w27211. 2020.

